# The Greek version of the Richards-Campbell Sleep Questionnaire: Reliability and validity assessment

**DOI:** 10.1101/2023.06.24.23291857

**Authors:** Christina-Athanasia Sampani, Marios Charalampopoulos, Panagiota Triantafyllaki, Christos Triantafyllou, Dimitrios Papageorgiou

**Affiliations:** General Hospital of Athens “G. Gennimatas”, Athens, Greece; Medecins Sans Frontieres-Greece; General Hospital of Athens “Evangelismos”, Athens, Greece; National and Kapodistrian University of Athens, School of Health Science, Department of Nursing, Goudi Athens, Greece; Director of “ICU Follow-Up Care Lab”, School of Health and Care Sciences, Department of Nursing, University of West Attica

**Author notes:** **Corresponding author:** Christos Triantafyllou, Papadiamantopoulou 123, Goudi Athens, PC 11527. On Behalf: ICU Follow-Up Care Lab, Department of Nursing, University of West Attica, Athens, Greece.

**Keywords:** Sleep in intensive care unit, polysomnography, Richards-Campbell Sleep Questionnaire, interventions on sleep

## Abstract

**Aim:** To translate the RCSQ into Greek and to determine the reliability and validity of the Greek version of the RCSQ as a measure of sleep among ICU patients in a Greek hospital.

**Methods:** This study investigated the night-time sleep of 50 patients admitted to the general ICU of General Hospital of Athens “G. Gennimatas” between January 2022– March 2023.

The RCSQ questionnaire was used for the present study with the written permission of its author. All guidelines for translating and adapting psychometric scales were followed for translating the RCSQ into Greek. Demographic-clinical characteristics (sex, age, days in hospital, days on mechanical ventilation, days on spontaneous breathing, whether they underwent tracheotomy and the type of tracheotomy, and whether they received a mild sedation formulation to promote sleep) were collected from the participants. Reliability was tested using Cronbach’s alpha coefficient.

**Results:** The Cronbach’s correlation coefficient *α* was calculated 0.906 revealing excellent reliability of the RCSQ. No question removal significantly increased the coefficient value.

**Conclusion:** The Greek version of the Richards-Campbell Sleep Questionnaire could be used as an alternative to polysomnography when assessing sleep quality in intensive care unit patients.

## Introduction

Although the function of sleep is not clearly understood, it is generally accepted that sleep is necessary for the maintenance of good health and the lack of sleep increases illness.

Sleep occurs in a deepening cycle of four stages followed by rapid-eye-movement sleep (REM sleep). Stage 1 is a transitional stage between wakefulness and sleep and is not considered to be real sleep. The sleeper is still aware of his surroundings and is easily aroused. Stage 2 is a slightly deeper stage of sleep, the sleeper is unaware of his surroundings but is still easily aroused. Stages 3 and 4, also called deep sleep, since this is the deepest stage of sleep, and it is very difficult to wake the sleeper during this period.^1^

Sleep quality in intensive care unit (ICU) patients is one of the problems that arise during their hospitalization. Approximately 51% of patients report a change in their sleep habits during hospitalization and after discharge from the ICU. The lack of sleep that many ICU patients suffer from can lead to confusion, more difficult rehabilitation, and prolonged morbidity.^2^

The ICU environment, mechanical ventilation, medication, and disease severity itself have been reported as important factors that disrupt sleep. The secretion of the sleep hormone, melatonin, which expresses circadian rhythmicity, was found to abolish or delay the sleep phase in critically ill patients.^3^

The causes of sleep fragmentation are not completely specific. However, many studies have been conducted that have demonstrated several. The patients treated in an ICU are severely ill and require monitoring of vital functions and care around the clock. A study in which the patients were asked to indicate what disturbed their sleep during their stay in the ICU demonstrated that monitoring procedures were felt to be the most disturbing.^4^

Monitoring and therapeutic procedures were shown by Meyer et al. (1994) to disturb patients on average about once per hour continuously throughout the day. According to several studies conducted on the subject matter, it has been found that noise levels in ICUs often surpass established norms. This is usually caused by both medical equipment and conversation among staff at high levels.

High noise levels can disrupt the overall quality of sleep regardless of whether or not an individual wakes up.

Strong lighting has a detrimental effect on sleep due to its impact on the foundation of the circadian rhythm, which is light. The reason for this is that intense light causes a halt in the production of melatonin. Melatonin plays an important part in the regulation of the sleep-waking cycle and in setting the biological clock. It is probable that the patients’ sleep-waking cycle is disrupted in an ICU where the lights are on at night and where there are loud noises, and the patients are disturbed by care routines at night as well.^5^§

The assessment of sleep in an ICU also remains a challenge. While polysomnography (PSG) remains the preferred method for evaluating sleep quality due to its reliability, it may not be practical for all situations due to its high cost as well as requiring the patient’s compliance during lengthy monitoring periods. However, actigraphy provides an objective way to measure sleep in the ICU. A wristwatch-like device called actigraphy is used for sleep analysis since it’s less invasive than PSG. Actigraphy may not be the most precise measure for detecting sleep stages, but PSG can provide that information.^6^

## Background

In subjective methodologies, the patients themselves describe the quality and quantity of their own sleep, usually with the aid of a questionnaire. The advantage of these methods is that it takes relatively little time to collect information on patients’ sleep and process the data. The disadvantages are that a number of different instruments/questionnaires used in studies of sleep consist of a very large number of questions. There are also difficulties encountered in securing validity and reliability. Severely ill ICU patients may not cope with answering such a lot of questions. Moreover, many of these patients have impaired cognition owing to the effects of the medication, sensory over- or under-stimulation, and some patients are unconscious.

As lack of sleep is a major problem for ICU patients, it is an important task for nurses to promote patients’ sleep by means of various care interventions. In order to be able to evaluate the effect of different interventions, a simple instrument is needed to help the nurse assess the patient’s sleep.^7^

### The RCSQ Instrument

Richards et al. (2000) developed and validated the Richards Campbell Sleep Questionnaire (RCSQ), a 5-item visual analog scale with an optional noise item, for critically ill patients assessing the following domains: sleep depth, falling asleep, number of awakenings, percentage of time awake and quality of sleep. The 6th Item should be scored individually. It was not part of the original RCSQ but can be used as a measure of noise.^8^

The scores for each item can be expressed on a scale from 0, marking the poorest-quality sleep, to 100, indicating optimum sleep. The use of the RCSQ has been described as easy and not time-consuming.^9^

The Richards-Campbell Sleep Questionnaire (RCSQ) shows a good correlation with PSG.

The RCSQ has been translated into other languages, including Spanish by Nicolas et al., (2008), Swedish by Frisk & Nordstrom, (2003), Japanese by Tsuruta, Yamamoto, & Fujita, (2017), and German by Krotsetis, Richards, Behncke, & Kopke, (2017). Evaluating sleep using the RCSQ is the most clinically relevant as the research in the lack for versatility of sleep evaluation using PSG.^1,7,10,11^

Since its development, the RCSQ has been widely used in international practice and sleep research to determine individually perceived quality of sleep in critically ill patients.^11^

## Methods

### Aim

The aim of the present study is to translate the RCSQ into Greek and to determine the reliability and validity of the Greek version of The RCSQ as a measure of sleep among ICU patients in a Greek hospital.

### Participants

This study investigated the night-time sleep of 50 patients admitted to the general ICU of G.N.A GENNIMATAS Hospital between January 2022– March 2023. The ICU had 17 beds in total and 1:2 nurse-to-patient ratio.

The included patients were over 16 years of age, with or without the need for mechanical ventilation and with hemodynamic stability. Patients younger than 16 years of age, patients with hemodynamic instability, sedated patients, patients with a history of sleep-disordered breathing (e.g., sleep apnea syndrome), patients with chronic neuromuscular disease, patients with psychiatric disease, patients with a previous history of sleep pathology, alcohol addiction or illicit drug abuse, and patients with a history of cognitive dysfunction (defined as the presence or history of dementia).

Data logs were anonymous and confidential. Data collection and processing will only be performed after the informed and free consent of the patient. The principle of integrity and confidentiality was ensured. The data in the questionnaires were coded, anonymous, and confidential, and a code number was written without any reference to the patient’s name.

### Description of RCSQ

The RCSQ questionnaire was used for the present study with the written permission of its author. All guidelines for translating and adapting psychometric scales (Wild et al., 2005) were followed for translating the RCSQ into Greek. Initially, the translation was carried out by two experienced and independent translators and the back-and-forth translation procedure was followed.^12^

The RCSQ scale is a short, self-report, 5-item questionnaire used to assess night sleep. Specifically, it assesses:

- sleep depth
- sleep latency (the time it takes to fall asleep)
- number of awakenings
- efficiency (percentage of time awake)
- sleep quality.

Each RCSQ item is scored on a visual analog scale ranging from 0 mm to 100 mm, with higher scores representing better sleep. The average score of the five items is known as the “total score” and represents the overall perception of sleep. As was done in previous studies, our questionnaire also included a sixth item assessing perceived noise at night (range: 0 mm for “very quiet” to 100 mm for “very noisy”).

Finally, demographic-clinical characteristics (sex, age, days in the hospital, days on mechanical ventilation, days on spontaneous breathing, whether they underwent a tracheotomy and the type of tracheotomy, and whether they received a mild sedation formulation to promote sleep) were collected from the participants.

### Statistical analysis

Data analysis was performed with the statistical package SPSS version 22. Mean (mean) and standard deviation (SD) were used to present the quantitative variables while absolute (N) and relative frequencies (%) were used to present the poetic variables. Normality tests were performed using Kolmogorov-Smirnov test. Reliability testing of the RCSQ scale was performed with Cronbach’s correlation coefficient *α*. Index values greater than 0.7 or 0.8 are usually considered satisfactory. To investigate the differences between the RCSQ and the demographic-clinical characteristics of the participants, parametric t-test and Mann-Whitney non-parametric tests were used. Pearson’s correlation coefficient (r) parametric and Spearman’s correlation coefficient (r) nonparametric were used to investigate the associations between two quantitative variables. Finally, a factor analysis was performed on the RCSQ scale. The level of statistical significance was set at p<0.05.

## Results

### Demographic characteristics

Of the 50 patients who took part in the study, 32 were men, and 18 were women. The mean age of the patients was 56.6 years (SD=17.9). The mean duration of hospitalization was 25 (SD=28.1) days, the mean duration of mechanical ventilation was 20.3 (SD=25.0) days, and the mean duration of spontaneous respiration was 4.9 days (SD=17.1; Table 1).

**Table 1.**
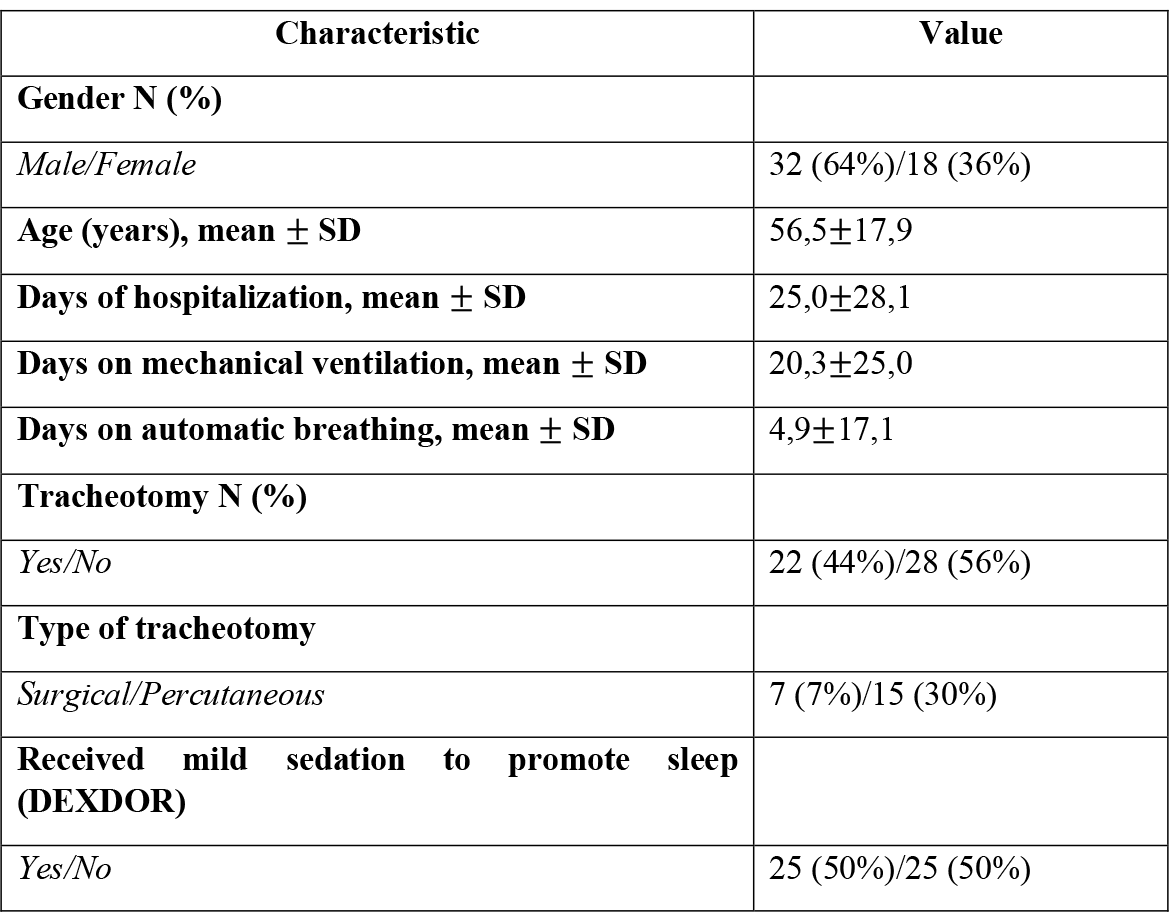
Characteristics of participants (n=50)

### Cronbach’s *α* coefficient

The Cronbach’s correlation coefficient *α* was calculated at 0.906 revealing excellent reliability of the RCSQ. No question removal significantly increased the coefficient value.

### Characteristics of RCSQ

The overall mean score of the RCSQ scale was 52.8 (SD=15.41) with a range of 12 to 78. Patients were divided into four groups according to the RCSQ scale score: patients with very poor sleep, patients with poor sleep, patients with good sleep and patients with very good sleep. 60% of the participants had a “good sleep” while 8% had a “very poor sleep” (Figure 1). **Table 2** shows that patients rated noise levels moderately (mean=57.0 SD=18.43).

**Figure 1.**
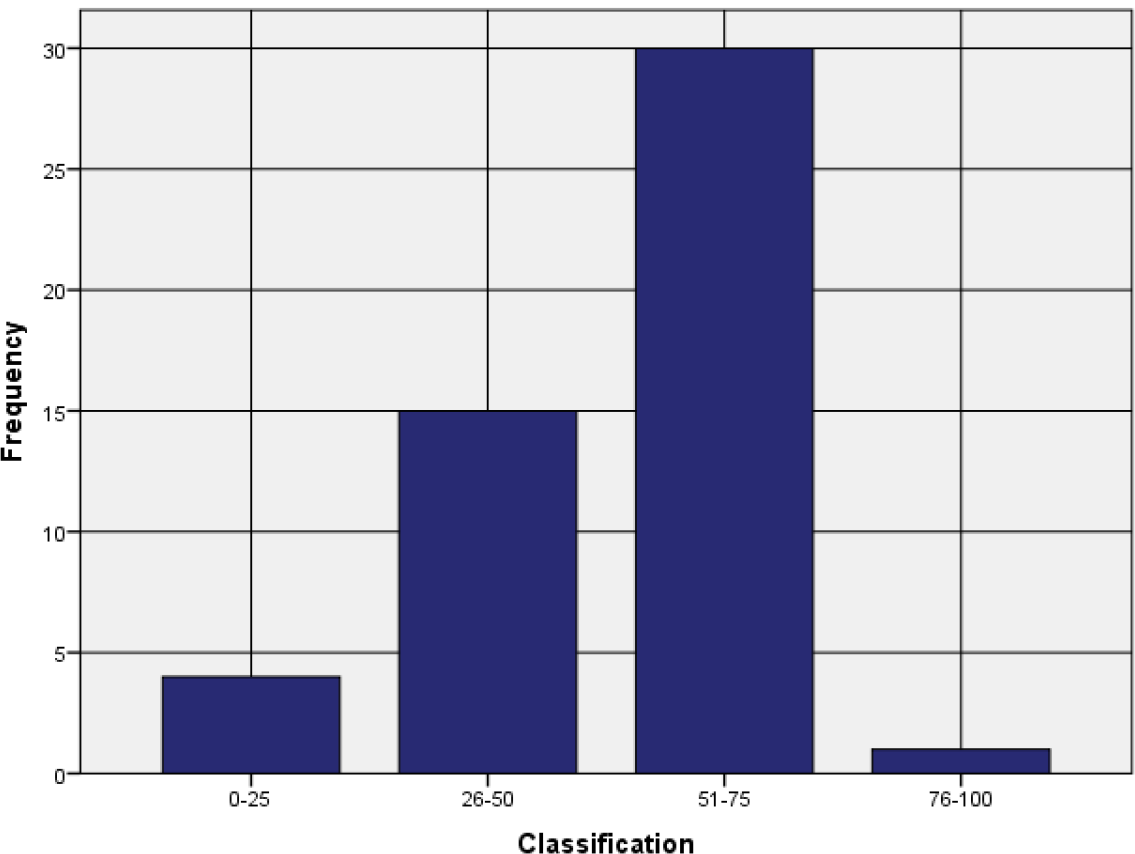
The groups of patients

**Table 2.**
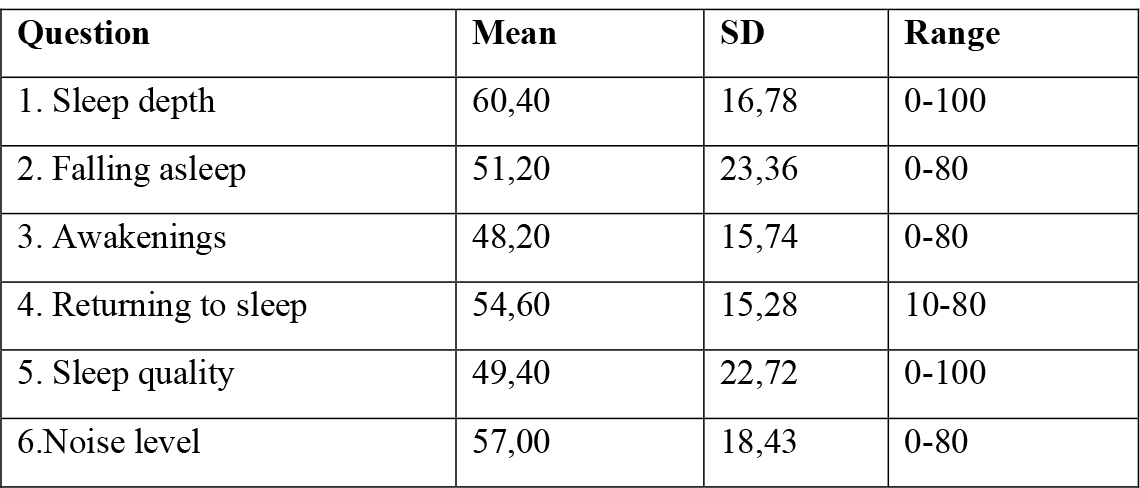
RCSQ Questionnaire

| Question | Mean | SD | Range |
| --- | --- | --- | --- |
| 1. Sleep depth | 60,40 | 16,78 | 0-100 |
| 2. Falling asleep | 51,20 | 23,36 | 0-80 |
| 3. Awakenings | 48,20 | 15,74 | 0-80 |
| 4. Returning to sleep | 54,60 | 15,28 | 10-80 |
| 5. Sleep quality | 49,40 | 22,72 | 0-100 |
| 6.Noise level | 57,00 | 18,43 | 0-80 |

### Comparisons of RCSQ and noise level

The analysis of gender and taking mild sedation for sleep promotion showed no statistically significant correlations for total score and noise level (p>0.05; Table 3).

**Table 3.** Comparisons of RCSQ and noise level with gender and received mild sedation to promote sleep

|  | **Gender** |  |  |
| --- | --- | --- | --- |
|  | **Male** | **Female** | **p** |
| **Total RCSQ** | 48,88±15,61 | 49,00±15,79 | 0,768 |
| **Noise level** | 53,75±17,08 | 51,67±27,87 | 0,725 |
|  | **Received mild sedation to promote sleep** |  |  |
|  | **Yes** | **No** | **p** |
| **Total** | 47,57±16,50 | 51,25±13,60 | 0,503 |
| **Noise level** | 52,86±18,16 | 53,75±23,87 | 0,721 |

### Correlations of RCSQ and noise level

The Age and the days of spontaneous breathing did not show a statistically significant correlation with the total RCSQ score (p>0.05). On the contrary, there was a negative statistically significant correlation between the total RCSQ score and patients’ hospital days (p=-0.495, p<0.05). Furthermore, there was a negative statistically significant correlation between the total RCSQ score with days on mechanical ventilation of patients (p=-0.474, p<0.05).

Days of mechanical ventilation did not affect the noise levels (p=-0.121, p>0.05). On the other hand, days in the hospital and days on mechanical ventilation of patients negatively affect noise levels (p=-0.465, p<0.05 and p=-0.451, p<0.05, respectively). Age appeared to be weakly and negatively correlated with noise levels (p=-0.289, p<0.05).

Finally, a statistically significant positive correlation was observed between the total RCSQ score and noise level (p=0.931, p<0.05).

### Factor analysis

Factor analysis showed that the Simple Moving Average (SMA) equals 0.824 and the statistical function of Bartlett’s sphericity test equals χ^2=230.054 and p<0.05 therefore, the scale items are uncorrelated with each other and suitable to be analyzed through factor analysis. The analysis resulted in a statistically significant factor explaining 70% of the total variability. The loading coefficients of all items were greater than the allowable limit.

## Discussion

Sleep disruption in the acute hospital setting has been reported widely in the literature. Patients have attributed sleep disruption to environmental factors, symptom management, and nursing interventions.^13^

Around 30% of patients are dissatisfied with their night’s rest, which nurses often fail to recognize. As the primary caregivers to patients in the hospital environment, nurses are strategically placed to assess and promote their patients’ sleep. However, it is possible that awareness of the health impacts of good sleep is suboptimal by nursing staff in the acute hospital environment, leading in turn to a lack of emphasis on procedure with regard to patient sleep promotion. One of the barriers to sleep promotion is the lack of a standardized tool to assess sleep.^14^

Routine use of an easy-to-use, brief sleep measurement tool has the potential to help nurses identify sleep-related issues and communicate this in a concise manner. Clarification of these issues could provide an impetus for improved nursing assessment, which should, in turn, lead to the implementation of appropriate nursing interventions that promote better sleep for patients. For clinical purposes, there is a need for a validated, brief sleep assessment tool for hospitalized patients.

Many studies reported the use of the RCSQ, which was originally developed as a five-item scale for patients to report on their previous night’s sleep in the critical care environment. According to the developer, Richards et al., 2000, items correlated with polysomnography in the domains of sleep onset, sleep depth and awakenings and total sleep time.^15^

The RCSQ takes approximately two minutes to complete. Item values are summated and divided by five providing a mean score reflecting the patient’s perception of their sleep. Thus far, the RCSQ has had limited application beyond the critical care setting where validation studies have been conducted.^15,16,7^

However, a feasibility study of elderly hospitalized patients compared the RCSQ with actigraphy and noted a moderate correlation (r = .57, p = <0.001).

The RCSQ has been used largely in critical care settings. Measured against polysomnography, it displayed positive psychometric properties in reliability and correlating with those polysomnographic measures capturing some of the domains of sleep quality in terms of sleep onset, awakenings and depth of sleep. Its ease of scoring, brevity and non-burdensome time commitment from the patient and/or carer make it an attractive option for evaluation of patient sleep assessment in an acute hospital ward.

One of the aims of the present study was to give an account of ICU patients’ perception of their sleep. The fact that a large number of the patients were not able to answer questions about their sleep is, of course, a problem, but one that is unavoidable if you want to study the perceptions of a group of individuals consisting of severely ill intensive care patients. However, it must be considered important to know how these patients experienced their sleep.

This finding is relatively well in line with the study by Nicolas et al. (2008), where one hundred and four surgical patients were recruited for the study. Patients completed the Richards-Campbell Sleep Questionnaire. The total mean score of sleep on the first post-operative night was 51-42 mm, 28% of patients had good sleep, 46% a regular sleep, and 26% a bad sleep. The sleep profile of these patients has been characterized by the patients having a light sleep, with frequent awakenings and generally little difficulty in going back to sleep after the awakenings.^1^

Freedman et al. (1999), in their study, measured the patients’ ratings of how they had slept the night before. Some stated that they had not slept at all, while others stated that they had slept very well. The total sleep score ranged between 0 and 97 (mean 45.5). Nine patients (29%) scored a total sleep score of 25 or below (i.e., very poor sleep), and eight (26%) a total sleep score of over 75 (i.e., very good sleep). Cronbach’s alpha for the five questions about sleep was 0.92. The results show very wide-ranging variation in the patient’s experience of their sleep.^4^

Similar findings had the study conducted by H. Murata et al. (2019). The Richards-Campbell Sleep Questionnaire was initially translated **into Japanese** using the back-translation method. Validity was evaluated by determining the association between sleep efficiency, measured using simplified polysomnography, and the total score on the Japanese version of the Richards-Campbell Sleep Questionnaire. Reliability was tested using Cronbach’s alpha coefficient. Thirty-three patients were included in the analysis. After excluding four patients with subsyndromal delirium, the Pearson correlation coefficient was 0.602 (*p* = 0.001). Cronbach’s alpha coefficient was 0.911. The Japanese version of the Richards-Campbell Sleep Questionnaire could be used as an alternative to polysomnography when assessing sleep quality in intensive care unit patients.^10^

In the settings of the study of S Krotsetis et al. (2017), the RCSQ was translated following established methodological standards. Data were collected cross-sectionally in a sample of 51 patients at three intensive care units at a university hospital in Germany. The German version of the RCSQ showed an overall internal consistency (Cronbach’s alpha) of 0⋅88. The mean of the RSCQ in the sample was 47⋅00 (SD ± 27⋅57). Depth of sleep was rated the lowest, and falling asleep again the highest of the RCSQ sleep items.^11^

Moreover, Hana Locihová et al. (2019) translated the RCSQ according to the translation and cultural adaptation manual. The quality of sleep was assessed using the Czech version of the RCSQ. The sample consisted of 105 patients hospitalized in an interdisciplinary intensive care unit. The internal consistency (Cronbach’s *α*) of the Czech version of the RCSQ is 0.89. There was no statistically significant relationship (p < 0.05) between sleep quality and selected variables: age (F = 0.1; p = 0.736), gender (F = 0; p = 0.929), and type of admission (F = 1.8; p = 0.183). This study demonstrates that the Czech version of the RCSQ is rated as a reliable tool and can be used to subjectively assess sleep quality in critically ill patients.^8^

In another study, Chen, Li-xia, et al. (2019) translated the original RCSQ into Chinese and then back-translated it into English to ensure its accuracy of the translation. Internal consistency, discrimination validity, and construct validity of the RCSQ were examined in 150 critically ill patients. The convergent validity of the RCSQ was evaluated in 44 of 150 critically ill patients, and data from the RCSQ were compared with those of the Chinese version of St Mary’s Hospital Sleep Questionnaire (SMHSQ). Cronbach’s *α* of the RCSQ was 0.923; thus, it showed high reliability. The psychometric properties of the RCSQ suggest its utility in critically ill patients.^17^ The Swedish version of the RCSQ, evaluated by Frisk and Nordström (2003), had a Cronbach’s alpha of 0.92 in 31 alert patients in a 6-bed surgical ICU in Sweden.^7^ The Spanish version by Nicolás et al. (2008), tested on n=104 non-mechanically ventilated patients in a 16-bed surgical ICU in Spain, had an alpha of 0.89.^1^

Our study, investigating a heterogeneous patient sample, importantly contributes to the applicability of the RCSQ in various intensive care settings.

The fifty included patients were over 16 years of age, with or without the need for mechanical ventilation and with hemodynamic stability. Cronbach’s correlation coefficient *α* was calculated to be 0.906, revealing excellent reliability of the RCSQ. No question removal significantly increased the coefficient value. On the contrary, there was a negative statistically significant correlation of the total RCSQ score with the patients’ hospitalization days (p=-0.495, p<0.05). Furthermore, there was a negative statistically significant correlation between the total RCSQ score with days on mechanical ventilation of patients (p=-0.474, p<0.05).

Assessment of patient sleep should be made using a valid and reliable instrument. The Richards– Campbell Sleep Questionnaire has the greatest potential for use in the acute hospital setting. It is brief, easy to use, and understandable and has demonstrated reliability and validity in the capture sleep domains to provide a global assessment of sleep. However, it should be subject to further testing.

### Limitations

To address the current study adequately, it is necessary to tackle several limitations. A limitation that may be considered is the variability of the patient’s individual histories regarding the use of medication or other sleep disorders. Although efforts will be made to exclude patients with these potential factors, the results of the current study may underestimate or overestimate the association between the outcomes. Also, the small sample of patients as well as the access to the ICU in a hospital, can be considered as possible limitations of the study.

Hence it is clear that generalizing the outcomes to people in critical conditions, such as patients with multiorgan system failure, is constrained, and the validation of RCSQ among various patient groups poses a challenge.

### Implication for practice

- Sleep disorders are frequently seen in all intensive care unit patients (compared to the general ward patient population).
- Sleep deprivation and disturbed sleep quality have clear and straightforward consequences for a patient’s level of distress.
- Many fail to utilize tools that could help assess the overall quality of their sleep by nurses.
- The assessment of sleep patterns using the Greek version of the RCSQ questionnaire proves to be a dependable approach.
- Patient experience with sleep quality in the ICU is subjective and personal.
- Interventions improving the quality of sleep could affect the global critical care outcome of intensive care unit survivors and should be a part of good quality clinical practice in the future.

## Data Availability

All data produced in the present study are available upon reasonable request to the authors

## References

1. Nicolas, Ana, et al. “Perception of night-time sleep by surgical patients in an intensive care unit.” Nursing in critical care 13.1 (2008): 25–33.

2. Al Mutair, Abbas, et al. “Sleep Deprivation Etiologies Among Patients in the Intensive Care Unit: Literature Review.” Dimensions of Critical Care Nursing 39.4 (2020): 203–210.

3. Boyko, Yuliya, Poul Jennum, and Palle Toft. “Sleep quality and circadian rhythm disruption in the intensive care unit: a review.” Nature and science of sleep 9 (2017): 277.

4. Freedman, Neil S., Natalie Kotzer, and Richard J. Schwab. “Patient perception of sleep quality and etiology of sleep disruption in the intensive care unit.” American journal of respiratory and critical care medicine 159.4 (1999): 1155–1162.

5. Meyer, Thomas J., et al. “Adverse environmental conditions in the respiratory and medical ICU settings.” Chest 105.4 (1994): 1211–1216.

6. Elliott, R., McKinley, S., Cistulli, P., & Fien, M. (2013). Characterisation of sleep in intensive care using 24 hour polysomnography: An ob- servational study. Critical Care, 17(2), R46. https://doi.org/10.1186/cc12565

7. Frisk, Ulla, and Gun Nordström. “Patients’ sleep in an intensive care unit—patients’ and nurses’ perception.” Intensive and Critical Care Nursing 19.6 (2003): 342–349.

8. Locihová, Hana, et al. “Sleep quality assessment in intensive care: actigraphy vs. Richards-Campbell sleep questionnaire.” Sleep Science 134 (2020): 235.

9. Hoey, Lynn M., Paul Fulbrook, and James A. Douglas. “Sleep assessment of hospitalised patients: a literature review.” International journal of nursing studies 51.9 (2014): 1281–1288.

10. Murata, Hiroaki, et al. “The Japanese version of the Richards-Campbell Sleep Questionnaire: Reliability and validity assessment.” Nursing Open 6.3 (2019): 808–814.

11. Krotsetis, Susanne, et al. “The reliability of the German version of the Richards Campbell Sleep Questionnaire.” Nursing in critical care 22.4 (2017): 247–252.

12. Wild, Diane, et al. “Principles of good practice for the translation and cultural adaptation process for patient-reported outcomes (PRO) measures: report of the ISPOR task force for translation and cultural adaptation.” Value in health 8.2 (2005): 94–104.

13. Hultman, Todd, et al. “Exploring the sleep experience of hospitalized adult patients.” Creative nursing 18.3 (2012): 135–139.

14. Yilmaz, Meryem, Yazile Sayin, and Hesna Gurler. “Sleep quality of hospitalized patients in surgical units.” Nursing forum. Vol. 47. No. 3. Malden, USA: Blackwell Publishing Inc, 2012.

15. Richards, Kathy C., Patricia S. O’Sullivan, and Robin L. Phillips. “Measurement of sleep in critically ill patients.” Journal of nursing measurement 8.2 (2000): 131–144.

16. Kamdar, Biren B., Dale M. Needham, and Nancy A. Collop. “Sleep deprivation in critical illness: its role in physical and psychological recovery.” Journal of intensive care medicine 27.2 (2012): 97–111.

17. Chen, Li-xia, et al. “Richards-Campbell sleep questionnaire: psychometric properties of Chinese critically ill patients.” Nursing in critical care 24.6 2019): 362–368.

